# Bridging the Nutrition Education Gap: From Theory to Practice- A Scalable Model for Nutrition Practicums in Medical Training

**DOI:** 10.1101/2025.02.18.25322448

**Authors:** Anjum John, V.R Reshma, Khadija El-Hazimy

## Abstract

**Background:** With Kerala’s rising burden of non-communicable diseases (NCDs), it is crucial to strengthen nutrition education for medical students to equip future physicians’ with the ability to provide evidence-based dietary guidance. This study examined the effectiveness of incorporating practical demonstrations into the nutrition curriculum of a private medical college in Kerala, Southern India. This aims to enhance student engagement, deepen understanding of dietetic principles through practical application, and improve confidence in providing dietary counseling.

**Methods:** The study incorporated practical demonstrations such as meal planning, dietary assessments, and food label interpretation into the nutrition curriculum for third year medical students. Student feedback was collected through open-ended discussions and reflective exercises to assess their engagement and perceptions of the perceived learning outcomes.

**Results:** Qualitative feedback indicated that students found practical demonstrations highly engaging, improving their confidence in applying nutritional knowledge. Thematic analysis identified key benefits such as enhanced experiential learning, increased ability to interpret nutrition labels, and improved patient counseling skills. However, challenges included limited time for hands-on activities, variability in prior nutrition knowledge, and limited faculty expertise in nutrition education.

**Conclusions:** Practical demonstrations are an effective strategy for integrating nutrition education into medical curricula in Kerala as underscored by this study. By aligning with the National Medical Commission’s emphasis on integrated, application-based learning, and Kerala’s goals of addressing its rising burden of non-communicable diseases linked to malnutrition, this study offers a scalable model to strengthen clinical nutrition education, ultimately contributing to better public health outcomes nationwide.

## Background

Food is essential for survival, and medicine has long recognized the importance of diet and nutrition for healthy living. It has become evident with time that diet and nutrition interventions go a long way in mitigating and preventing lifestyle-related diseases such as obesity, diabetes, and cardiovascular conditions.(1) Medical education had traditionally placed *limited* emphasis on nutrition, often relegating it as a supplementary topic with limited time allotment, rather than placing it as a central pillar of clinical practice.(2,3) Medical teachers are uniquely positioned to enhance public health by the exclusive opportunity to educate future doctors early in their training on the importance of diet and nutrition in patient care, shaping their lifelong clinical practices.(4)There is a critical need to enhance knowledge of nutrition in medical teachers and students *alike*.(1,5)

Current curricula often fall short of the required practical, hands-on experience necessary for medical students to translate theoretical knowledge of diet and nutrition into effective dietary guidance for patients.(6,7) This gap is compounded by a deficiency of integrated teaching methods with nutritional practicums that blend scientific principles into real-world applications. The gap is particularly evident in regions where formal nutrition education is not emphasized in medical curricula.

Nutrition science is a domain where misinformation and fads thrive, highlighting the need for students to rely on evidence-based research to become well-informed educators. (8)To address these challenges, there is a growing interest in incorporating practical demonstration techniques—such as interactive workshops, cooking classes, and dietary assessment exercises—that equip students with hands-on experience and critical thinking skills to navigate and counter nutrition myths effectively.(4,8)These methods can not only foster a deeper understanding of nutrition but also empower medical students to confidently discuss and recommend dietary modifications for patients as well as apply them to their own lives.(8,9)

Though medical curricula worldwide mandate nutrition education, there is a lack of standardization regarding its placement and mode of delivery within these programs. This inconsistency creates challenges to educationists in ensuring that students acquire the knowledge and skills necessary for effective patient care in disease management. (10) In France, nutrition is incorporated into the Endocrinology curriculum of medical schools, following up on foundational courses in physiology, biochemistry, or cell biology. (11)In India, nutrition is primarily part of the Community Medicine curricula, instructed by medical faculty who often lack specialized training in dietetics.(12) Nutrition and medicine are taught in the United States (US) and the United Kingdom (UK) as a unified science rather than compartmentally.(13,14)

Absence of standardized epistemological approaches to nutritional pedagogy has resulted in different methods being used for teaching nutrition, aligning with the specific needs of the curricula, patient and community demands, or national priorities. One of these methods is the inclusion of culinary medicine programs like Teaching Kitchens(15), or Health Meets Food, (16,17)developed within individual medical schools in the United States (US), which emphasise hands-on experiential learning to effectively transmit knowledge. As culinary kitchen programs collaborate with communities, hospitals, schools, medical professionals, and the food industry, they help students develop a broader perspective on the profound impact of food on both patients and personal health. Reading food labels on packaged food is another skill that will serve medical students in good stead. Interpretation of nutrition labels is a simple method by which students can understand and act on claims made by the packaged food industry. This is particularly important because research suggests a strong association between higher consumption of packaged foods, obesity markers and many non-communicable diseases.(18,19)

Understanding the nutritional needs of the population through diet surveys and diet histories, are community-based population approaches that have been used in nutrition education.(12) These help students make recommendations for diet changes and understand the importance of nutrition management in patient care. Studies have also shown that assessing the knowledge and skills of medical students through incorporating questions on nutrition within formal assessments can help improve their communication, diagnostic and referral skills.(8,12)

Building on this, the current study aims to explore the potential of integrating practical demonstrations of dietary and nutritional techniques into medical education, focusing on their impact on students’ knowledge, hands-on skills, and self-perceived confidence in applying nutrition principles in clinical practice. Key competencies aimed for included taking a basic food history, reading, and analysing food labels, and delivering effective and empathetic nutritional counselling using evidence- based guidelines, which are culturally sensitive and devoid of bias and judgments. (21, 22) By examining the effectiveness of these teaching strategies, this research seeks to offer insights into how medical curricula can evolve to better equip future healthcare providers with the skills essential to address the dietary needs of their patients and community.

This study is among the few, conducted in this part of the world, which includes Kerala, where nutrition education is traditionally underrepresented in medical training. By highlighting an innovative, experiential learning model, this research offers valuable insights into how competency-based medical training (CBME) can be enhanced to better equip future healthcare professionals with essential nutrition counseling skills among others, thus addressing a well-documented, yet unresolved gap in their practice.

With the burden of diet-related non-communicable diseases and malnutrition posing a significant public health, economic, and social challenge, medical professionals must be adequately trained to provide effective nutritional counseling as part of patient management.

### Theory

According to the Kolb’s Experiential Learning Theory (ELT), for effective learning to take place, a cyclical process of engaging in a specific activity leads to a concrete experience.

When the learner reflects on that concreted experience, gains insights, and applies what is learnt to new situations, his or her understanding and skills are reinforced. This research incorporates practical demonstration techniques into nutrition and dietetic learning. Hands-on activities can foster reflective thinking, encourage learners to derive meaningful insights, and can deepen the understanding of the concepts learned. In the future, students can apply the knowledge learned in real world situations, through active experimentation. (20)

### Theoretical Framework

The theoretical framework integrates Experiential Learning Theory with the Constructivist Learning Theory, where students build knowledge actively through practical experiences which foster retention of concepts learned and create deep understanding of the subject.

Active participation of students in interactive activities like meal planning, peer critiquing of diet plans, enhances confidence, applying knowledge into practice helps internalize concepts and build practical skills, and qualitative feedback aligns with reflective observation and helps students improve their learning outcomes.

### Conceptual Framework

The conceptual framework outlines the relationship between the teaching method (practical demonstration techniques with hands on learning as an independent variable), with student engagement through participation, and reflection of insights from qualitative feedbacks and, as mediating variables, and learning outcomes like improved understanding of dietetics principles, self-professed ability to apply knowledge in clinical scenarios, and confidence in addressing diet-related patient issues serving as dependent variables. (20)

### Methodology

#### Study Design and Setting

This educational innovation study was conducted as part of a structured nutrition course for third-year medical students.

The intervention integrated practical demonstrations, including meal planning, dietary counseling techniques, and nutritional assessments, within a competency-based medical education curriculum (CBME). Student feedback was collected through open-ended discussions and reflective exercises. The research utilised a single-group observational study design. As part of the year 3 curriculum for medical students, 15-18 hours were designated for diet and nutrition classes. The four-day practical nutrition course was part of the mandated curriculum of the National Medical Council of India. All students had completed their basic science course which included Anatomy, Physiology, and Biochemistry

#### Inclusion and Exclusion Criteria of the participants

Thirty third-year MBBS (Bachelor of Medicine and Surgery degree course) students were enrolled in a four-day practical diet and nutrition course. Recruitment for student participants took place in the classroom setting using the convenient sampling method. No consent was taken from the students at this time.

### Study Design

This study employed a mixed-methods educational evaluation with a post-course assessment approach, incorporating qualitative feedback and rubric-based quantitative assessments to explore student learning outcomes.

The study was conducted in two phases: one, an intervention phase and second, the assessment and feedback phase. The intervention phase was conducted on the cohort of 30 medical students.

The assessment and feedback phase utilized qualitative and quantitative data, with student feedback collected through open-ended discussions and reflective exercises to explore learning outcomes in the absence of a comprehensive structured evaluation. The study included post-tests and rubric-based assessments conducted during exams after the course, but did not incorporate standardized pre-tests or other systematic methods for measuring learning gains over time.

### Interventions

The interventions were carried out in stages.

1. **Theory Sessions**: Assistant professors from the Department of Community Medicine instructed the students via PowerPoint presentations. Topics covered included principles of nutrition, dietary requirements for various populations, how to read food labels, and the role of nutrition in managing specific health conditions.
2. Practicums

a. **Interactive Sessions**: Students participated in interactive sessions that included discussions on dietary requirements for different populations- both healthy and diseased- with specific health conditions attributable to malnutrition. Tools like the 24-hour recall and Food Frequency Questionnaire(21) were used to conduct nutritional assessments. These were done as part of the Family Adoption Programme(22), in the field area of the Community Medicine Department.
b. **Creating Sample Meal plans:** Practical sessions that encouraged real world application of concepts through creating sample meal plans for different populations- both real and hypothetical- and discussing the preparation of balanced meals designed to provide a well-rounded learning experience that bridged the gap between theoretical knowledge and clinical application. Food ingredients for activities were provided by the teaching faculty while available departmental equipment was used for the practicums.
c. **Learning simulations using hypothetical scenarios**: Using case scenarios, students were taught meal planning for diverse populations and dietary counselling for specific health conditions. They were used as input for detailed simulation outputs, aligned with Kolb’s Experiential Learning Theory to capture hands-on learning outcomes.
d. **Evaluation of Student Competencies in Nutrition Assessment and Counseling**: Course instructors assessed student performance using established rubrics with predefined criteria. Students were evaluated on their ability to apply nutritional screening tools (e.g., 24-hour recall, Food Frequency Questionnaire) to assess nutritional status in community settings. They also created tailored nutritional intervention plans based on hypothetical patient cases, incorporating evidence-based dietary guidelines (e.g., low-sodium diet for hypertension). Empathy in communication was subjectively assessed through case scenarios, where students developed culturally sensitive meal plans supported by current nutritional guidelines and research.
e. **Qualitative Feedback on Learning and Application**: Feedback was gathered through open-ended group discussions and self-reported observations, where all students collectively reflected on their engagement, learning experiences, and confidence in applying nutrition knowledge, allowing students to express their perspectives on the effectiveness of the course.
f. Literature Review: Building on the qualitative feedback, researchers conducted a literature review on practical techniques in medical nutrition education to further contextualize student experiences and identify effective teaching methods and implementation barriers. What began as a teaching and learning exercise gradually evolved into a research endeavour, as student reflections highlighted valuable insights worth documenting. This review not only informed the development of questionnaires for a subsequent study on hands-on nutrition education in subsequent classes but also laid the groundwork for a separate narrative review exploring best practices in the field.
g. Thematic Analysis: The qualitative data were analysed to identify common themes related to experiential learning benefits and challenges.

### Ethical Considerations

This study began as an educational innovation with formal assessments, and where qualitative feedback was collected to enhance teaching strategies. Ethical transparency was maintained by informing students about the purpose of feedback collection, and no identifying student data were used in the analysis. Since the course was conducted without an initial intent for research, consent was not obtained at the time of data collection. After recognizing the research value of the collected data, ethical approval was sought and granted by the Institutional Review Board (Approval No. PIMSRC/E1/388A/156/2024) for its retrospective use. The requirement for consent was waived, as the data was not traceable to individual participants, and any publications presenting only aggregated, non-identifiable results.

## Results

This study highlights participants’ knowledge, perceptions, and confidence in nutrition as assessed through post-test evaluations and qualitative feedback, providing insights into the effectiveness of the intervention.

As shown in Tables 1-3, the post-test assessments revealed notable outcomes across key domains, reinforcing the effectiveness of the hands-on learning approach.

**Table 1:** Distribution of Student Scores in Post-Class Nutrition Theory Assessment (Total of 30 Marks)

| Question | 0-6 Marks | 7-12 Marks | 13-18 Marks | 19-24 Marks | 25-30 Marks | p-value |
| --- | --- | --- | --- | --- | --- | --- |
| Basic human nutrition | 0 (0%) | 1 (3%) | 5 (17%) | 15 (50%) | 9 (30%) | 0.000 |
| Dietary guidelines | 0 (0%) | 3 (10%) | 6 (20%) | 13 (43%) | 8 (27%) | 0.000 |
| Patient's nutritional status | 0 (0%) | 1 (3%) | 6 (20%) | 15 (50%) | 8 (27%) | 0.000 |
| Role of diet in chronic diseases | 0 (0%) | 3 (10%) | 5 (17%) | 17 (57%) | 6 (20%) | 0.000 |
| Nutritional needs of population | 5 (17%) | 8 (10%) | 5 (17%) | 6 (20%) | 2 (7%) | 0.000 |

Table 1 presents the distribution of student scores in the post-class nutrition theory assessment, which had a total possible score of 30 marks. Across all five assessed domains, the majority of students scored in the higher ranges, indicating a strong understanding of the covered topics. In the Basic Human Nutrition, Patient’s Nutritional Status domain, Dietary Guidelines, and in the Role of Diet in Chronic Diseases sections, the majority scored between 19-24 marks, with a small percentage obtaining the highest scores. However, performance in the Nutritional Needs of the Population category was notably lower, with only 7% achieving scores between 25-30 marks, and a considerable proportion (17%) scoring in the lowest range (0-6 marks). A statistically significant difference was observed across all domains (p = 0.000), suggesting meaningful variations in student performance across the different aspects of nutrition theory.

Additionally, students expressed confidence in discussing nutrition, making dietary recommendations, and integrating nutrition into clinical practice. Compared to the traditional lecture-based instruction in the first part of the course, which focused primarily on theoretical knowledge, students perceived the experiential, skills-based learning approach as more effective in building practical competencies. They reported that the interactive nature of the course helped bridge the gap between theory and real-world application, enhancing their ability to assess and counsel patients on nutrition-related issues. Table 2 summarizes the post-intervention assessment of medical students’ confidence in various aspects of nutrition knowledge, as evaluated by teachers. The majority of students were rated as confident across all domains, with a smaller proportion remaining neutral or not confident.

**Table 2:**
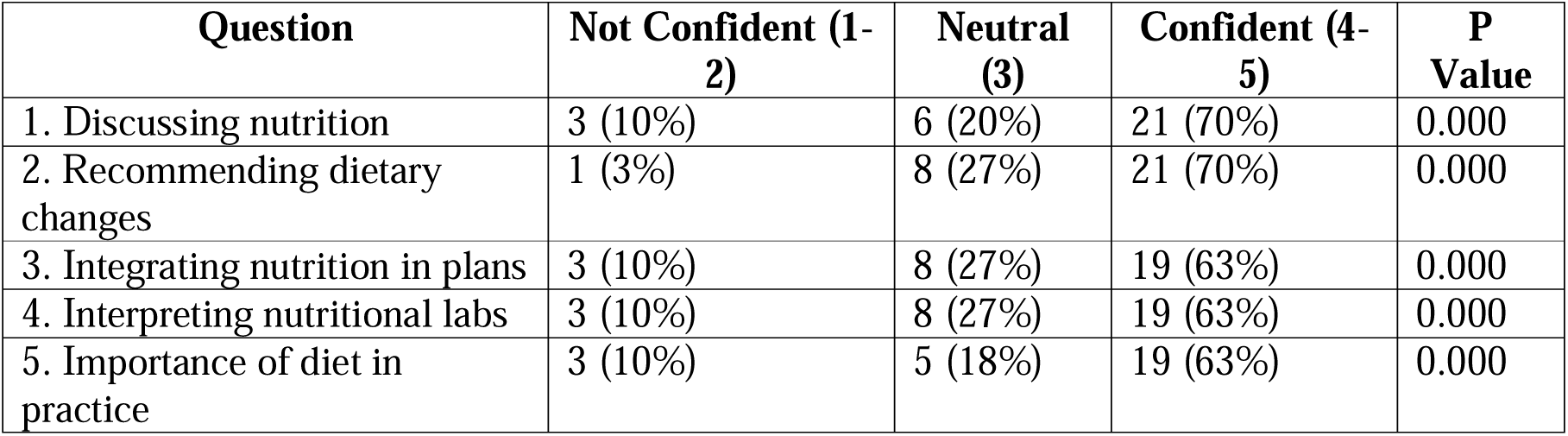
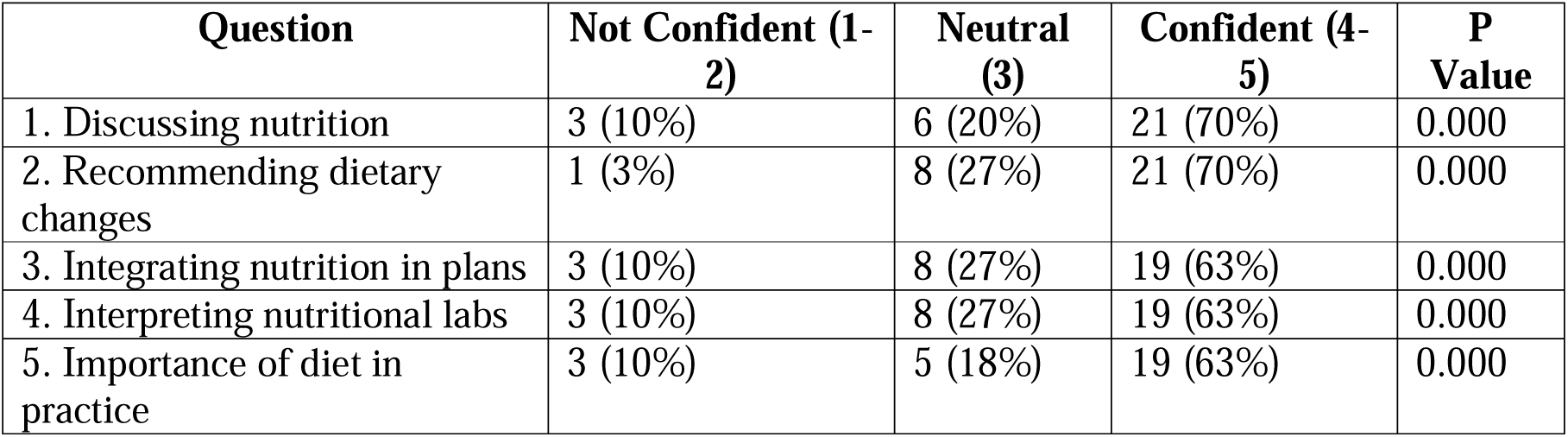
Post-Intervention Assessment of Medical Students’ Confidence in Nutrition Knowledge: Teacher Evaluations.

### Post-Test Assessments (Table 3)

Table 3 presents the teacher-assessed performance of medical students in key competencies related to nutritional evaluation and counseling. The majority of students demonstrated proficiency across all assessed areas, with most scoring in the Excellent (90-100%) or Good (75-89%) categories. Competencies such as correct use of screening tools, realistic and culturally sensitive meal planning, and clear and empathetic communication had the highest proportion of students rated as excellent. A smaller proportion of students fell into the Satisfactory (50-74%) category, while very few required improvements.

**Table 3:**
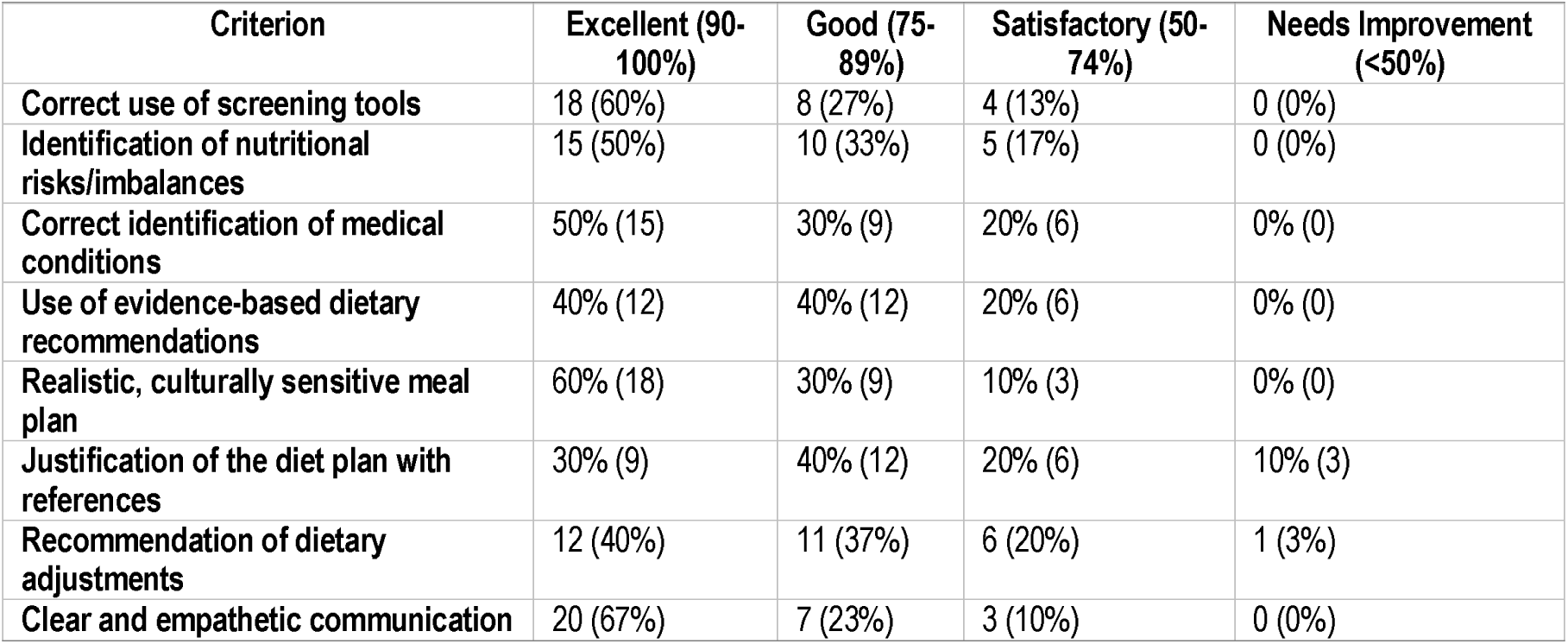
Performance assessment of key competencies in nutritional evaluation and counselling (teacher assessments)

The exercise on identifying nutritional risks through case scenarios revealed variability in student performance. While 50% of students demonstrated strong analytical skills, others showed only partial competence, missing key details in dietary assessments. This highlighted the need for improved training.

When asked to recommend dietary adjustments based on identified risks, 40% of students excelled, providing comprehensive, evidence-based recommendations. An equal proportion performed well but lacked depth in justifying their dietary plans.

Empathy in nutrition counselling, assessed through hypothetical case scenarios, showed the strongest student performance, with 67% demonstrating clear, patient-centered communication. However, some students needed improvement in conveying dietary advice in an accessible and culturally sensitive manner. These results highlight the effectiveness of the intervention in enhancing students’ ability to apply nutrition knowledge in clinical settings However, targeted training for lower-performing students could further enhance outcomes.

Table 4: Bridging Theory with Practice: Insights from Qualitative Feedback: Medical Students’ Perspectives on Nutrition Education (Reflections)

**Table 4:**
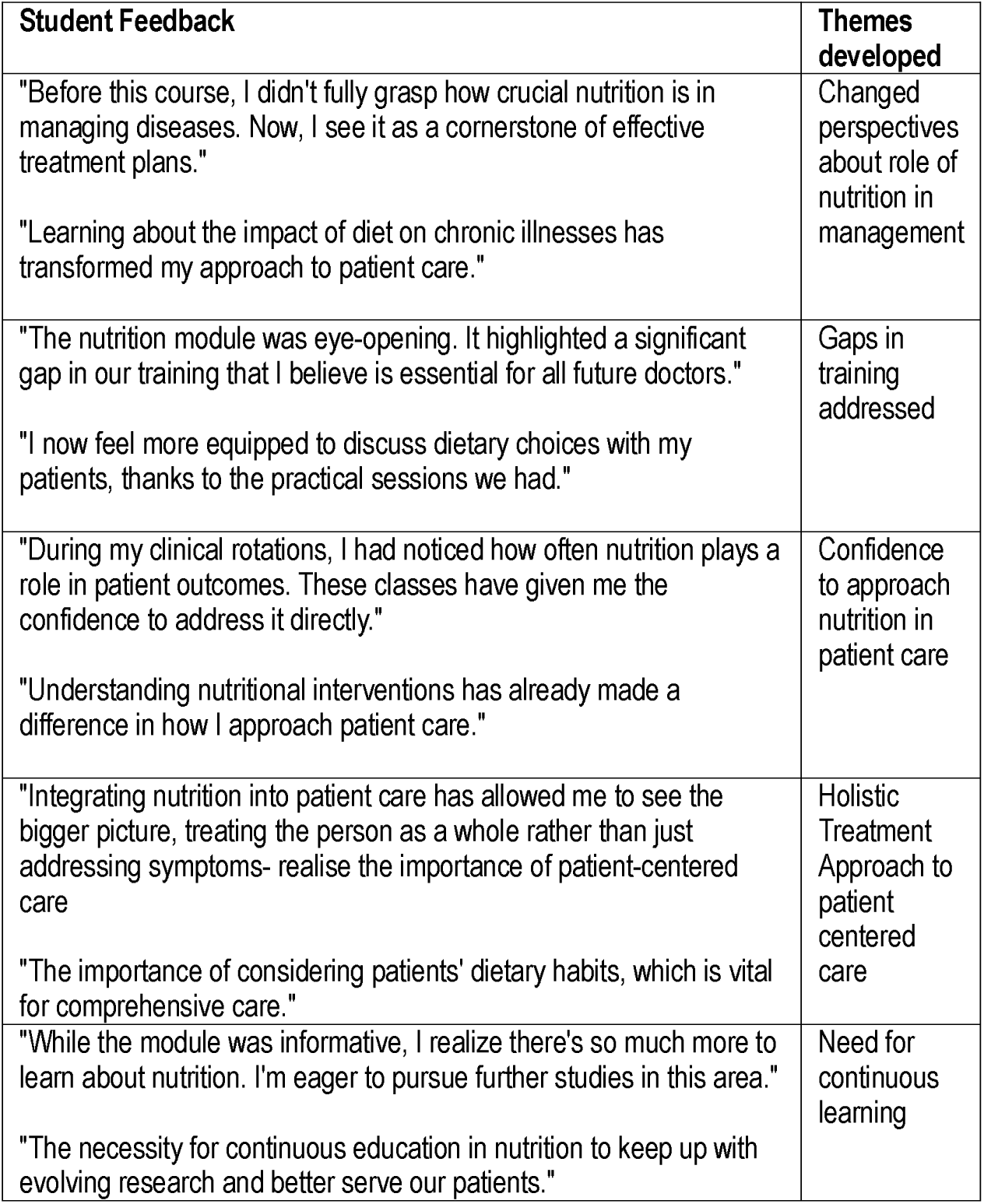
Medical Students’ Perspectives on Nutrition Education (Reflections)

The qualitative component of the study captured students’ reflections on their learning experiences, engagement, and confidence in applying nutrition knowledge in real-world contexts. Compared to lecture-based teaching, students reflected that hands-on, practical demonstrations were far more engaging and effective.

Students highlighted key themes regarding the relevance and impact of the nutrition module. Many students reported a *changed perspective on the role of nutrition* in disease management, recognizing it as a fundamental aspect of effective treatment plans. They expressed they felt more prepared to discuss dietary choices with patients and thus the course addressed *gaps in their training*.

A notable outcome was the *increased confidence in incorporating nutrition into patient care*, as students acknowledged how nutritional interventions influenced clinical decision-making. Additionally, students emphasized the importance of a *holistic, patient-centered approach*, integrating dietary habits into comprehensive care rather than focusing solely on disease symptoms. Finally, several students expressed a *desire for continuous learning*, recognizing the need to stay updated on evolving nutrition research to enhance patient care.

Students found interactive activities—such as meal planning, food label interpretation, and real-world nutritional assessments—highly engaging. These activities fostered deeper participation and helped reinforce clinical nutrition concepts. Many students expressed that traditional lecture-based teaching had failed to equip them with practical skills, whereas experiential learning significantly improved their confidence in providing nutrition-related counselling.

Students reported that experiential learning methods, including case scenario analysis and problem-solving exercises, helped them develop critical thinking and decision-making skills in clinical nutrition. The integration of hands-on activities with theoretical concepts enhanced their ability to apply nutrition knowledge effectively in patient care.

Moreover, the collaborative nature of practical sessions prepared students for real-world, multidisciplinary healthcare environments. They appreciated opportunities to work in teams, simulate patient interactions, and practice evidence-based nutrition counselling.

Time constraints and varying levels of prior nutrition knowledge were noted as barriers to deeper engagement.

These findings underscore the impact of the nutrition teaching methods in improving students’ perceptions, confidence, and preparedness in applying nutrition knowledge in clinical practice.

## Discussion

The findings highlight the effectiveness of practical demonstrations in medical nutrition education.

### Overview of Findings

This study, conducted among thirty medical students, evaluated the outcomes of various practical nutrition demonstration interventions. Post-course assessments and qualitative feedback indicated that students gained valuable practical skills and confidence in applying nutrition knowledge. However, areas such as “confidence in applying nutrition knowledge” and “perceived relevance in clinical practice” showed variability, suggesting opportunities for refining the curriculum to better support skill development.

### Global Perspectives on Nutrition Education

Several international studies emphasize the critical role of nutrition education, particularly hands-on training, in equipping medical students to counsel patients, evaluate dietary patterns to recommend appropriate dietary changes. A 2023 study of French and American medical schools identified gaps in nutrition curricula and underscored the value of multidisciplinary, experiential approaches. Social scientists have recommended pedagogical strategies tailored to nutrition education to bridge these gaps.(11)

In India, the National Medical Council (NMC) recommends incorporating nutritional interventions for health promotion and rehabilitation through the Attitude, Ethics, and Communication (AETCOM) module. However, nutrition education is not formally integrated into the medical curriculum and is typically addressed only when time permits. Few published studies from India focus on medical nutrition education, and motivated faculty often resort to innovative teaching methods to compensate for the lack of structured training.(23,24)

### Historical Context

In 2014, Dr. Shankar advocated for formal nutrition education in India’s medical curriculum, proposing a tailored “Nutrition in Medicine” module inspired by the University of North Carolina’s program. He emphasized the need to adapt nutrition education to India’s cultural diversity and integrate it into medical training to address the nation’s dual burden of over- and undernutrition. (25)

### Hands-On Approaches in Nutrition Education

Recognizing India’s rising burden of chronic diseases linked to poor diets, Gandhi et al. introduced a hands-on “Diet Demonstration Program,” inspired by Culinary Kitchen classes in the U.S. This program encouraged students to apply theoretical knowledge by cooking with locally sourced ingredients. Most participants reported enhanced understanding of nutritional practices and greater confidence in counseling patients. (26)

## Findings and Recommendations

The practical demonstration exercise showed that most students effectively used screening tools, such as 24-hour dietary recalls and food frequency questionnaires. However, significant performance disparities among student groups suggest the need for targeted training. Limited curriculum time constrained additional teaching, necessitating self-directed learning for underperforming students.

A 2019 systematic review by Crowley et al. highlighted global inadequacies in medical students’ nutrition knowledge, confidence, and skills, largely due to time constraints, lack of qualified educators, and logistical challenges in implementing nutrition curricula. These findings echo the gaps identified through this study.(27)

A study conducted in a neighbouring district, published in May 2024, showed that students recognized the importance of nutrition education for diet counseling. However, they found dietary surveys using the 24-hour recall method challenging in practical situations. (12)

## Comparisons with Lecture-Based Learning

Students perceived the hands-on, skills-based learning approach as significantly more effective than traditional lecture-based instruction. While theory classes provided foundational knowledge, students felt such knowledge gained lacked practical applications in clinical scenarios. The interactive nature of the practical sessions helped bridge the gap between theory and practice in the real world, reinforcing their ability to assess and counsel patients on nutrition-related issues. However, since no pre-tests were conducted, improvements in knowledge and confidence could only be inferred from post-course feedback rather than if they were direct comparisons with baseline levels.

## Empathy in Nutrition Counseling

The study revealed that most students demonstrated empathetic communication skills, crucial for patient-centered care. A 2024 scoping review by de Graaf et al. emphasized empathy as integral to effective nutritional counseling, improving patient adherence to dietary recommendations. Structured pedagogical strategies can further enhance students’ empathetic capacities. (28)

## Strengths of the study

This research emphasizes the crucial role of practical training in improving diet and nutrition education within medical curricula. Hands-on sessions would equip medical students with skills to identify dietary risks, formulate evidence-based dietary recommendations, and effectively utilize dietary screening tools.

Practical exercises helped students develop culturally relevant, evidence-based dietary plans and understand real-world challenges, such as preparing affordable, balanced meals.

However, variability in the quality of the evidence generated through student feedback and assessments, as well as inconsistencies in the organization of their dietary plans, highlight the need for additional training in systematically justifying nutritional decisions and structuring their findings effectively. Most students demonstrated a good understanding of evidence-based practices, as seen in the alignment of their plans with recommended dietary guidelines. Some students noted the challenges of preparing affordable and balanced meals, reflecting an awareness of real-world constraints. Understanding local ingredients and cooking practices helped students create culturally relevant dietary plans in hypothetical situations. The exercise helped students appreciate the importance of using scientific evidence to support dietary advice, although the level of thoroughness varied across distinct groups.

These discussions provided an open forum for students to share insights, challenges, and suggestions, fostering peer learning and deeper understanding, and the reflections therein captured their perceived strengths, challenges, and areas for improvement, particularly in using nutritional screening tools, developing intervention plans, and communicating dietary recommendations to patients.

The feedback provided insights into students’ experiences with nutritional screening tools, intervention planning, and communication of dietary recommendations, offering valuable input for refining teaching strategies. Additionally, these reflections gave the researchers a deeper understanding of student learning processes, helping identify key themes and gaps in student understanding, thus motivating them to refine instructional strategies to enhance experiential learning, in addition to giving them the idea to explore their findings in a research paper on nutrition education and experiential learning.

## Broader Integration in Medical Training

A sustained focus on nutrition education from the first year through internship is crucial. Innovative teaching methods, including case-based learning, role-playing, and culinary classes, can strengthen students’ theoretical knowledge and practical skills. While curriculum revisions by regulatory authorities like the NMC are underway, faculty and students must explore creative interim solutions.

Encouraging students to critique each other’s recommendations and communication skills may further help in cultivating the interpersonal skills essential for their future roles as practicing clinicians. (29)

Incorporating feedback from social scientists, dietitians, and medical educators can help students create balanced, realistic, and culturally sensitive dietary plans. Detailed scoring rubrics and peer review activities can enhance learning by clarifying expectations and fostering collaboration.(27)

The design of this research aligns with Kolb’s Experiential Learning Theory, as students engaged in practical activities such as dietary planning and food label interpretation, which helped them internalize what they learned. By connecting these activities to their theoretical nutrition knowledge, they developed a deeper understanding of dietetics. Students applied their knowledge to real-world clinical scenarios, such as counseling hypothetical patients, thus closing the loop of experiential learning. (20,30)

The connection with Constructivist Learning Theory is also evident, as students actively integrated prior knowledge with new experiences, allowing them to conceptualize and personalize their understanding of nutrition principles. This research could be further strengthened by incorporating hands-on practicum sessions involving food preparation or observation, which would highlight the role of social and cultural factors in nutrition, in accordance with Vygotsky’s Sociocultural Theory.(31)

The conceptual framework of this study demonstrates the interplay of theory, practicum, and student outcomes. Theoretical learning is integrated into the framework through faculty-led teaching of nutritional principles and practical tools like diet surveys. The learning process is reinforced through experiential activities such as case-based exercises, student feedback, and post-course assessments. The outcomes from this intervention include enhanced student confidence in applying nutritional principles, improved skills, and a better understanding of real-world dietary challenges.

The theoretical foundation, learning process, and student outcomes align with the medical curricular requirement of Competency-Based Medical Education (CBME). Educational innovation through experiential learning supports the CBME goal of producing practice-ready healthcare professionals. Post-course feedback and qualitative inputs from students align with Dewey’s philosophy of learning as a continuous reconstruction of experience.

Student reflections on their roles in addressing nutrition-related challenges helped them develop a deeper understanding of the subject. (32–34)

## Recommendations

To enhance nutrition education through medical curricula, this study recommends conducting training workshops focused on effective referencing and interdisciplinary collaboration, involving expertise from social scientists, dieticians, nutritionists, clinicians, and nursing professionals. The integration of artificial intelligence (AI) to generate culturally relevant evidence on nutritional practices should be explored, ensuring AI-generated content is supplemented with expert validation. Curricular innovations should prioritize innovative teaching strategies, such as simulations, virtual cooking sessions, hybrid modes of teaching, and telehealth dietary counseling, grounded in frameworks like Kolb’s Experiential Learning Model and the Five A’s Model for behaviour change.(20,35) Establishing a teaching kitchen within the Community Medicine laboratory can provide students with hands-on experience in meal planning and cooking, using locally sourced ingredients while addressing affordability and contextual, experiential learning.

Additionally, dedicated funding and structured frameworks with measurable competencies, such as case-based nutritional counseling and reflective writing assignments, should be implemented. The development of patient education materials can further enhance communication skills. Multidisciplinary training among departments like Physiology, Biochemistry, Pharmacology, and Community Medicine is crucial for a comprehensive approach to nutrition education, aligning with Vygotsky’s Sociocultural Theory. Encouraging international collaborations can promote shared learning opportunities to address similar healthcare challenges across different settings.

To overcome barriers in the learner-centric nutritional curriculum, faculty mentorship should be strengthened through workshops, bedside and outpatient learning, and online media for integrating evidence-based practices into curricula. Training faculty members in evidence-based practices and sociocultural dimensions of nutrition can facilitate team-based learning and ensure that future healthcare professionals are empathetic, competent, and equipped to address evolving nutrition-related challenges.(25,26)

## Barriers to Effective Nutrition Education

Barriers to effective nutrition education include overcrowded curricula, a shortage of medical faculty with specialized expertise, frequent changes in course instructors, limited resources, competing teaching priorities, and resistance to curricular change. Integrating nutrition education into already dense curricula may challenge the breadth of medical training.

Additionally, establishing infrastructure for experiential learning requires substantial financial and logistical investment, which resource-constrained institutions may struggle to sustain. Nutrition practices and dietary needs vary widely across regions and cultures, making it difficult to develop a standardized curriculum that maintains both local and global relevance. Moreover, the long-term impact of nutrition education on patient care and public health outcomes may take years to materialize, making its inclusion in medical curricula less immediately compelling compared to subjects with more direct and tangible clinical outcomes.

## Limitations

This study has several limitations that should be considered when interpreting the findings. First, the small sample size limits the generalizability of results to broader populations of medical students. Additionally, the study was conducted within a single institution, which may not reflect the diversity of nutrition education practices across different medical colleges regionally or nationally. The reliance on self-reported information introduces the potential for response bias, as participants may have overestimated their knowledge or confidence in alignment with socially desirable responses.

Furthermore, as only post-tests were conducted and no pre-tests were available for comparison, the study could not assess knowledge gains or confidence improvements relative to a baseline, or measure objective learning gains. While qualitative feedback from students provided insights into the perceived effectiveness of hands-on learning compared to traditional lecture-based instruction, objective measurements of long-term knowledge retention and clinical application were not included. The perspectives of faculty members and institutional leaders were also not explored, missing valuable insights into systemic challenges and opportunities for integrating nutrition education into medical curricula.

## Future Research

Future research should explore the impact of digital tools on personalized student learning outcomes and competency development in dietary counseling. The effectiveness of culturally tailored nutrition education modules, incorporating local dietary habits, cooking practices, and socioeconomic factors, should be evaluated. Additionally, interdisciplinary teaching approaches can be assessed for their ability to enhance students’ understanding of sociocultural, clinical, and community health aspects of nutrition. Future studies should explore integrating formative assessments to quantify learning outcomes more effectively.

Research should also examine the feasibility and effectiveness of establishing teaching kitchens in medical colleges as a means of promoting hands-on learning. New tools for assessing competencies gained through experiential learning, such as counseling skills and empathetic communication, should be developed and validated. Studies on faculty training workshops should focus on their long-term effects on student learning and professional practices, particularly regarding the integration of nutrition counseling into clinical practice and its influence on patient outcomes. Furthermore, cost-effectiveness and benefit analyses should be conducted to evaluate innovative teaching strategies, such as virtual cooking sessions and telehealth-based nutrition training, to ensure scalability and sustainability in resource-constrained settings.

## Conclusions

Kerala being a state facing rising rates of non-communicable diseases linked to poor dietary choices, and a growing epidemic of obesity and diabetes, this research will draw the attention of future healthcare professionals to practical nutrition education. The findings underscore the importance of experiential learning in improving students’ confidence and ability to integrate nutrition advice into patient care. Addressing gaps in nutrition education can enhance medical graduates’ capacity to provide evidence-based dietary counseling, contributing to better public health outcomes. As Kerala grapples with the dual burden of overnutrition and undernutrition, incorporating structured, hands-on nutrition education into medical training is an essential step toward improving healthcare delivery and chronic disease prevention.

In the broader Indian context, this study holds significance as medical education regulations evolve to prioritize competency-based medical education. The National Medical Commission (NMC)’s emphasis on integrated, practical, and application-based learning, align with the objectives of this study. By demonstrating the value of experiential learning in nutrition education, this research supports efforts to establish structured nutrition curricula within CBME, ensuring future physicians are well-equipped to address the country’s growing burden of nutrition-related diseases. This study presents a feasible, scalable model for integrating nutrition education into medical curricula, offering a framework that can be adapted across diverse healthcare education systems. Given the limited number of studies exploring practical interventions in medical nutrition education, this research fills a critical knowledge gap and provides an evidence-based approach to strengthening clinical nutrition training. Strengthening medical training in this domain is essential for improving dietary counseling practices and ultimately enhancing public health outcomes nationwide.

## Declarations

Ethics approval and consent to participate: After recognizing the research value of the collected data, ethical approval was sought and granted by the Institutional Review Board (Approval No. PIMSRC/E1/388A/156/2024) for its retrospective use. The requirement for consent was waived, as the data was not traceable to individual participants, and any publications presenting only aggregated, non-identifiable results.

## Consent for publication

Not applicable

## Availability of data and materials

All data generated or analysed during this study are included in this published article [and its supplementary information files].These include results of post class tests. No publicly archived datasets were analysed or generated during the study.

## Authors’ contributions

Dr. Anjum John taught the nutrition class for the medical students that semester. At the time of the class, I didn’t expect to write up a paper on this class but after teaching the class, I thought what I learnt through this teaching should be shared. Hence, the idea, the research protocol, the ethics approval submission, the data collection, analysis, result writing, manuscript preparation, journal submission were done under her supervision and guidance.

Ms. Reshma V.R, the biostatistician is currently unattached to any institution and moved to the United Arab Emirates at this time- she contributed by critiquing the proposal, helping with the data analysis, writing the results, reading the manuscript critically and providing general support for the project along with approval of the manuscript for final submission.

Ms. Khadija El-Hazimy is the Research and Data Specialist on the project; she provided all the references for the study and helped with the literature review and writing the discussion part of the manuscript, critiquing the manuscript and approval of the manuscript for submission.

## Data Availability

All data produced in the present work are contained in the manuscript.

## Acknowledgements

Dr. Felix Johns, Head of the Department of Community Medicine at Pushpagiri Medical College and Research Center approved the project Animal or human data or tissue : “Not applicable”

